# Contextualized Biomedical Language Processing Enhances ICU Survival Prediction

**DOI:** 10.1101/2024.12.20.24318979

**Authors:** Rui Chen, Yu Cai, Sitong Zhang, Zirong Huo, Mingming Song, Wenqing Li, Dongyan Yang, Seungyong Hwang, Ling Bai, Xi Zhang

## Abstract

**Background:** Cerebrospinal fluid (CSF) culture is the diagnostic gold standard for neuroinfectious diseases such as bacterial meningitis, but its sensitivity is limited and results are often delayed. Natural language processing (NLP) offers a powerful approach to extract meaningful clinical signals from unstructured data such as chief complaints and ICD notes. This study applies machine learning, including BioBERT-enhanced NLP models (not traditional TF-IDF approaches), to support early diagnosis and outcome prediction in ICU patients.

**Methods:** Training and validation datasets were derived from MIMIC-IV (internal) and MIMIC-III/eICU (external) databases. Fully connected neural network (FCNN) and other machine learning models were trained to predict CSF culture results using structured lab features. Labels were refined using clinical criteria to reduce false negatives. For ICU survival prediction, three multimodal deep learning architectures (mCNN, mFCNN, and mLSTM) were developed using two ICU survival cohorts with different inclusion criteria. The Strict ICU Survival Cohort included CSF culture results as an input feature, while the Lenient ICU Survival Cohort excluded this requirement, allowing for a broader patient population. In both cohorts, models integrated structured variables with unstructured text encoded by BioBERT, a deep contextual language model, rather than simpler methods like TF-IDF, effectively capturing clinical meaning from free-text ICD entries and chief complaints.

**Results:** For CSF culture prediction (training n = 9261), the FCNN model achieved the highest performance (AUROC = 0.853) in independent validation. For ICU survival prediction in the Strict ICU Survival Cohort (training n = 5,795), the mCNN model achieved an AUROC of 0.889 in external validation. In the expanded Lenient ICU Survival Cohort (n = 58,615), the same model achieved an AUROC of 0.974 and an AUPRC of 0.868 during external validation. During model training and development, the predictive performance declined when text features were excluded (AUROC from 0983 to 0.946) or when ICD entries were converted from free-text (BioBERT-encoded) to coded format (AUROC to 0.947).

**Conclusions:** Multimodal machine learning models, enhanced by advanced NLP through BioBERT embeddings of clinical free text, effectively predicted CSF culture results and ICU survival outcomes.

## Background

Neuroinfectious diseases, which include infections of the central nervous system such as bacterial and viral meningitis and viral encephalitis, can lead to significant complications if not diagnosed and treated promptly[1]. Compared with other types of neuroinfectious disease, bacterial meningitis (BM) typically has a rapid onset and high mortality rate which is largely attributed to delayed diagnosis and treatment[2]. In this study, we initially explored the utility of machine learning models in predicting cerebrospinal fluid (CSF) culture results and aiding BM diagnosis. However, the primary objective was to leverage these predictive features to develop robust models for forecasting intensive care unit (ICU) survival outcomes. By enhancing diagnostic accuracy and risk stratification, leveraging both structured lab results and unstructured clinical data, this approach has the potential to support early and effective clinical decision-making in critically ill patients.

The gold standard diagnosis of BM primarily relies on a combination of clinical findings and CSF analysis, with CSF culture serving as the definitive confirmation[3]. It is notable that CSF culture has time-consuming limitations and a high risk of false negatives[4, 5]. Because of these limitations, it is encouraged that CSF culture result should be processed alongside other lab test results and clinical evidences to improve diagnostic accuracy[6]. A major challenge lies in the overwhelming amount of clinical and laboratory data that clinicians must interpret in a short time frame. Machine learning models can help address this challenge by synthesizing complex datasets and highlighting relevant patterns, thereby supporting faster and more accurate clinical decision-making.

In addition to predicting CSF culture results, which are essential for diagnosing BM, assessing a patient’s survival offers clinically valuable insights into disease progression and outcomes. This task requires timely and precise decision-making from clinicians[7]. Real-time survival prediction models combine diverse patient information to estimate current condition and likely trajectory, helping to guide prediction and appropriate interventions[8]. While structured data such as laboratory results and vital signs are commonly used, unstructured text data also provide important clinical context. This includes chief complaints, clinical notes, and diagnostic impressions, which often contain nuanced information not captured in structured fields. One example is International Classification of Diseases (ICD) data, which can appear as standardized codes (such as “G00.9”) or in free-text form (such as “bacterial meningitis, unspecified”). The coded format supports categorization and billing, while the text format can offer additional context useful for clinical interpretation. This distinction is especially important in ICU settings, where patients are often unable to communicate their symptoms directly[9]. Recent advances in NLP, including transformer-based models like BERT (Bidirectional Encoder Representations from Transformers) and its biomedical adaptation BioBERT[10, 11] as well as models based on Neural Networks and Long Short-Term Memory (LSTM)[12, 13], have proven effective in capturing implicit relationships between terms and phrases related to patient prognosis by processing unstructured text data. Compared to traditional methods used for ICU prediction like TF-IDF[14], which rely on simple word frequency statistics and ignore context, BERT-based models provide deep contextualized language representations that better capture the meaning and clinical nuance of text. However, few studies have integrated NLP-derived insights with structured data for predicting outcomes such as survival in ICU patients. This represents a promising opportunity to develop multimodal models that improve predictive accuracy and clinical decision-making.

The primary objective of this study was to develop and evaluate machine learning models to predict ICU survival outcomes by identifying patterns in complex laboratory data and clinical evidence. During model development, we found that the likelihood of a positive CSF culture could also be predicted using structured lab data, suggesting that traditional culture results may be supplemented or even be replaced by model-driven insights. A key component of our approach was the integration of unstructured clinical text, such as ICD entries in free-text format as transformed by a BioBERT encoder. This NLP-enhanced feature set captured critical contextual information not present in structured data alone and proved essential for improving predictive accuracy. The resulting tools offer a culture-independent, data-driven solution to outcome prediction, supporting earlier and more informed decision-making in the ICU.

## Methods

### Data Source

This study utilized data from three publicly accessible critical care databases: the Medical Information Mart for Intensive Care-IV (MIMIC-IV v2.2), MIMIC-III (v1.4-subset), and the eICU Collaborative Research Database (eICU-CRD v2.0). MIMIC-IV contains structured (e.g., laboratory tests, vital signs) and unstructured data (e.g., clinical notes, discharge summaries) collected between 2008 and 2019 for 299,712 patients across 431,231 admissions and 73,181 ICU stays[15, 16]. MIMIC-III (v1.4-subset; 2001–2008) was used for independent external validation, and there is no patient overlap with the MIMIC-IV dataset[17]. Additionally, eICU-CRD (2014–2015), covering 200,859 ICU admissions across 208 U.S. hospitals was newly incorporated as another external validation dataset[18]. Database access required completion of the NIH “Protecting Human Research Participants” course (Certification ID: 10093146 and 12810933). This analysis adheres to the Reporting of Studies Conducted Using Observational Routinely Collected Health Data guidelines for administrative claims data[19].

### Textual Data Processing

Unstructured text features (chief complaint, ICD text, past medical history, history of present illness, allergies, and family history) were transformed into numerical vectors using BioBERT, a biomedical adaptation of BERT pre-trained on PubMed articles (Figure S1). Text tokenization converted inputs into semantic embeddings for integration with structured data. For MIMIC-III, where chief complaint data was absent, the biomedical-ner-all tool was used to extract equivalent information from clinical notes (Figure S2).

### Data Extraction and Feature Selection

For Culture Cohort and Survival_S Cohort, patients undergoing CSF bacterial culture tests were identified from MIMIC-IV, yielding 11892 cases. After excluding records missing essential biochemical tests, 11859 samples with 306 features and 2 label sets were retained (Figure. 1, Table S4). Structured data (n=300), including demographic, vital sign, blood test, and CSF biochemical results, underwent feature selection via the Least Absolute Shrinkage and Selection Operator (LASSO), reducing the dataset to the most relevant predictors[20]. Unstructured text data (n=6) was processed by extracting “Allergies,” “History of Present Illness,” “Past Medical History,” and “Family History” using regular expressions from discharge summaries, while “Chief Complaint” and ICD text data were directly accessed from database columns. For Survival_L Cohort, structured data extraction strictly followed Lim et al.’s criteria[8] (Table S4). Textual features (Chief Complaint, ICD text) were additionally extracted for model performance enhancement comparison.

**Figure 1.**
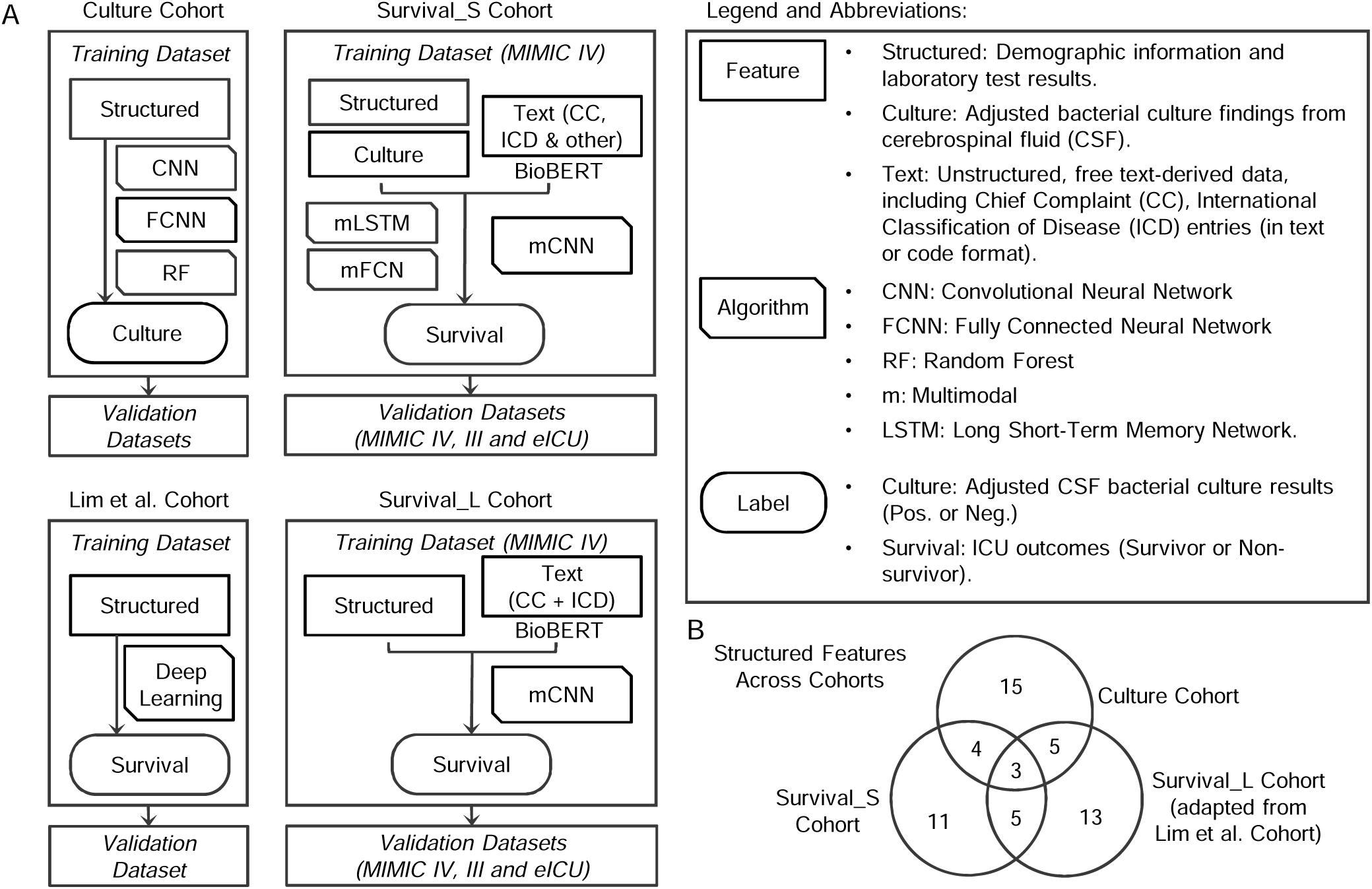
Study design and cohort setup. A. Overview of cohort construction, feature integration, model training, and validation strategies across three ICU cohorts evaluated in this study. Each cohort includes one training dataset (MIMIC-IV), one internal validation dataset (MIMIC-IV), and two external validation datasets (MIMIC-III and eICU). The Culture Cohort used selected structured data to predict cerebrospinal fluid CSF bacterial culture outcomes. Three models (CNN, FCNN, and RF), were evaluated with FCNN demonstrating the best performance. The Strict Survival (Survival_S) Cohort integrated optimized structured data, unstructured clinical text (chief complaints and ICD in text format etc.), and CSF culture outcomes to predict ICU survival outcomes. Among three multimodal models tested (mCNN, mFCNN, mLSTM), mCNN demonstrated the highest predictive performance. The Lenient Survival (Survival_L) Cohort adapted structured features from the reference Lim et al. Cohort and incorporated selected free-text data (chief complaints and ICD) without culture results, using the mCNN architecture selected from the Survival_S Cohort to assess ICU survival prediction. The Lim et al. Cohort represents the published ICU survival outcome model based solely on structured data, used here as a reference. The legend defines feature and algorithm abbreviations, and annotated prediction labels used for training. B. Venn diagram showing the overlap and unique structured features across the Culture, Survival_S, and Survival_L cohorts. Structured features for the Culture and Survival_S cohorts were identified via LASSO regression, while features for the Survival_L cohort were adapted directly from the reference Lim et al. study.

### Data Splitting and Preprocessing

The dataset was stratified into training, validation, and test sets (6:2:2 ratio) to ensure balanced class representation[21]. Missing values were imputed using a K-Nearest Neighbors Imputer (K = 5), and numerical features were normalized using the StandardScaler[22, 23]. Processed data was serialized for reproducibility. Text data was tokenized using BioBERT to create embeddings compatible with multimodal models.

### Model Development

Three multimodal neural network architectures (mFCN, mLSTM, mCNN) were developed to integrate structured and unstructured data. BioBERT embeddings of unstructured text were combined with structured data for model training: mFCN: Unstructured data embeddings were fed into a FCNN; mLSTM: Embeddings were processed by an LSTM network before integration with structured data; mCNN: Embeddings underwent a CNN-based transformation before combining with structured data. Baseline models trained exclusively on structured data were also developed for comparison.

### Data Analysis and Tools

Model performances were evaluated using accuracy, sensitivity, specificity, AUPRC and AUROC to comprehensively assess predictive performance for binary classification and survival prediction tasks. Distribution differences were assessed using boxplots and dotplots. Statistical significance was determined via two-sample t-tests or Mann-Whitney U test, and p values were adjusted for multiple comparisons with the Benjamini–Hochberg (BH) procedure to control the false discovery rate. Data processing and modeling were conducted using Python (v3.9.12) with libraries including pandas, numpy, and scikit-learn. TensorFlow and Keras were employed for neural network training, while BioBERT from Hugging Face handled text embedding. Due to the memory-intensive nature of the ‘labevents’ file form MIMIC datasets, systems with at least 64 GB RAM were required.

## Results

### Study Design and Multi-Cohort Modeling Framework

To achieve the primary objective of predicting ICU survival outcomes, we designed a multi-cohort modeling framework that also explored the feasibility of predicting CSF culture results using machine learning. Three ICU cohorts (Culture Cohort, Survival_S and Survival_L) were constructed to support stepwise model development, integration of structured and unstructured data, and external validation to predict ICU survivals (Figure 1A, Figure S1). All cohorts used MIMIC-IV for model training and internal validation, while MIMIC-III and eICU served as external validation datasets. Each cohort focused on a specific prediction task and included a tailored set of input features (structured, text and culture) and model architectures.

Structured features were either selected using LASSO regression (Figure S2) or adapted from a previously published ICU survival prediction study that supported real-time clinical decision-making (Figure 1B)[8]. Text features included chief complaints (CC), ICD entries in free-text format, and additional clinical text fields such as illness history, medical history, allergies, and family history. These unstructured features were iteratively optimized during model development to improve predictive performance.

### Predicting CSF Culture Results Using the Culture Cohort

The Culture Cohort consisted of 9,261 ICU encounters with available CSF data from MIMIC-IV, which were randomly split into 80% for training and 20% for testing (Figure S1). In clinical practice, CSF culture results are often interpreted in conjunction with other diagnostic indicators to address false negatives. For example, a CSF leukocyte count of ≥1000 cells/μL, particularly with a predominance of neutrophils, is commonly associated with bacterial meningitis and may be used to infer infection status in the absence of positive culture. Guided by this standard, we refined culture-positive cases using supporting evidence including elevated CSF leukocyte counts, high polymorphonuclear cell percentages, or chief complaint text explicitly referencing “bacterial meningitis”[24, 25]. This increased culture-positive cases from 245 to 404.

In the structured feature set, LASSO-optimized structured features included CSF-related (C-), blood test, and vital sign variables. CSF-derived features such as white blood cells (WBC), protein and glucose levels in CSF showed distinct distributions between culture-positive and culture-negative groups (Figure 2A). Baseline characteristics and statistical comparisons further highlighted significant group differences (Table 1 & S1). Three models (CNN, FCNN, RF) were trained to predict CSF culture status. The CNN model performed best (AUROC 0.813) in the initial cohort, with consistent trends in external validation (AUROC 0.769) (data not shown). Re-training the models with the updated labels led to improved performance. The FCNN model achieved the highest performance with an AUROC of 0.891, accuracy of 0.867, and sensitivity of 0.833 (Figures 2B & 2C), validating the utility of refined labeling in data-driven infection prediction. External validation on MIMIC-III (n = 1618) and eICU (n = 65) remained consistent, with AUROCs of 0.853 and 0.849, respectively, supporting the generalizability of the refined model.

**Figure 2.**
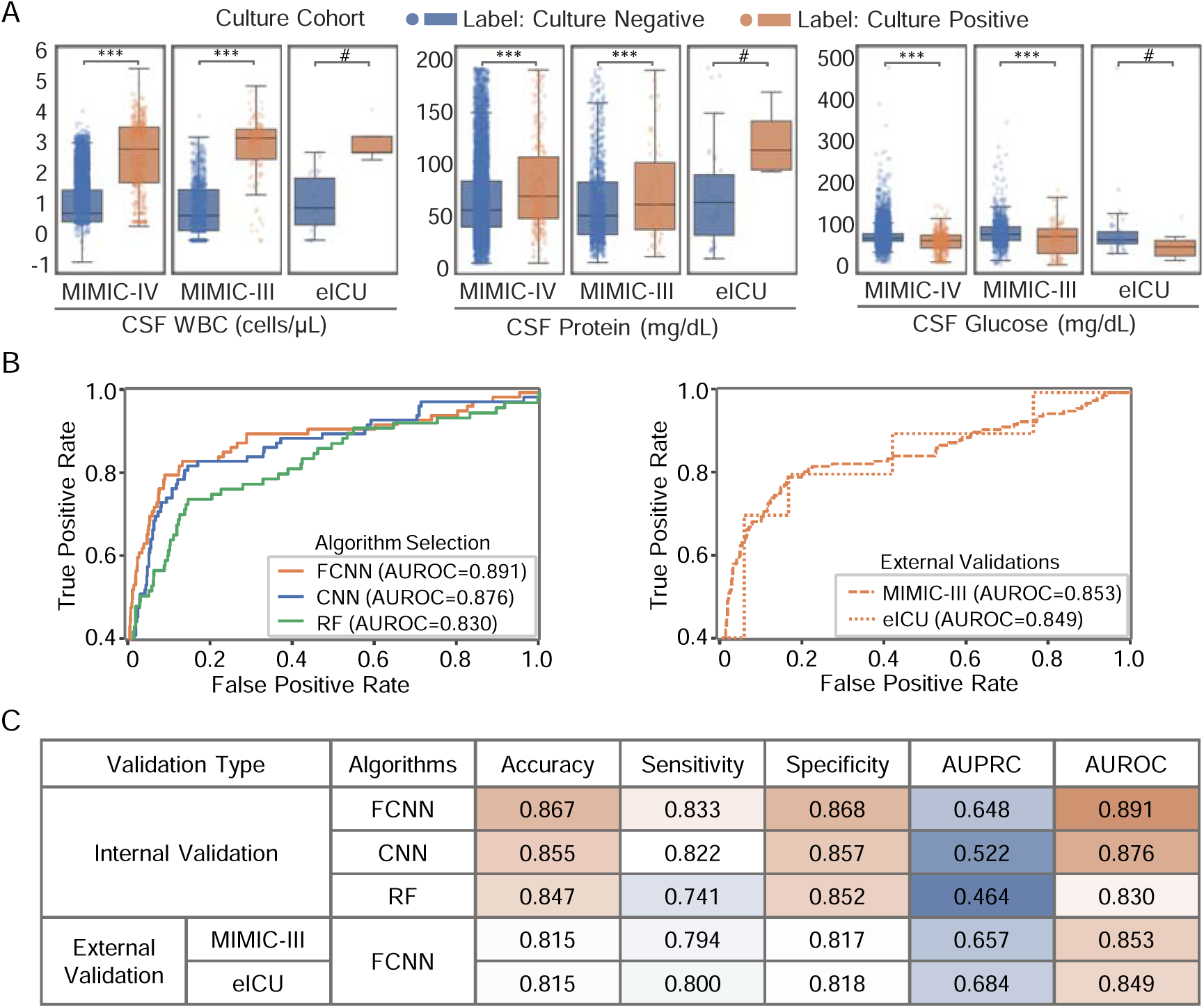
Model Development and Evaluation for Predicting CSF Culture Results in the Culture Cohort. A. Three representative structured features are plotted and compared between culture-positive and culture-negative groups across the MIMIC-IV, MIMIC-III, and eICU databases. B. ROC curves showing internal model performance (MIMIC-IV) across three algorithms (CNN, FCNN, RF), and external validation performance of the optimized FCNN model in MIMIC-III and eICU datasets. C. Summary table of performance metrics across internal and external validation datasets. CNN: convolutional neural network. FCNN: fully connected neural network. RF: random forest.

**Table 1.** Baseline Characteristics and Mean Comparisons of the Culture Cohort (MIMIC-IV)

| Features | Mean_neg | Mean_pos | Count_neg | Count_pos | P(adj) |
| --- | --- | --- | --- | --- | --- |
| C-Polys | 15.04 | 56.41 | 8733 | 390 | 6.92E-190 |
| C-Lymphocytes | 59.65 | 27.64 | 8733 | 390 | 1.56E-87 |
| C-WBC | 35.69 | 2446.68 | 8730 | 390 | 1.80E-79 |
| C-Protein | 64.69 | 251.49 | 8596 | 379 | 9.57E-56 |
| C-Glucose | 72.49 | 58.55 | 8494 | 366 | 7.15E-23 |
| Platelets | 238.5 | 279.96 | 8810 | 402 | 2.10E-12 |
| Hemoglobin | 11.78 | 11.01 | 8812 | 402 | 8.63E-11 |
| Albumin | 3.74 | 3.31 | 2757 | 95 | 5.71E-09 |
| Lymphocytes | 20.89 | 14.6 | 5388 | 160 | 6.90E-07 |
| Glucose | 121.69 | 132.48 | 8445 | 397 | 3.31E-05 |
| Pain | 4.34 | 5.8 | 2302 | 110 | 0.00189 |
| Potassium | 4.04 | 3.94 | 8635 | 399 | 0.00189 |
| C-Macrophage | 15.41 | 10.16 | 1876 | 103 | 0.00997 |
| Immature granulocytes | 0.59 | 0.8 | 1474 | 54 | 0.01940 |
| Monocyte count | 0.67 | 0.81 | 1818 | 69 | 0.03564 |
| Creatinine | 1.03 | 0.92 | 8706 | 400 | 0.05488 |
| Acuity | 2.12 | 2.02 | 2781 | 158 | 0.11806 |
| Sodium | 138.92 | 138.54 | 8636 | 399 | 0.11806 |
| Icterus index | 0.86 | 0.73 | 2463 | 126 | 0.15829 |
| RDW-SD | 47.48 | 48.57 | 3178 | 160 | 0.16049 |
| MCHC | 33.42 | 33.31 | 8810 | 402 | 0.21942 |
| LDH | 331.76 | 378.04 | 2190 | 72 | 0.53745 |
| Respiratory rate | 17.62 | 17.75 | 2422 | 131 | 0.71085 |
| pO2 | 125.26 | 121.6 | 1350 | 126 | 0.73687 |
| PTT | 30.45 | 30.6 | 4989 | 254 | 0.86292 |
| Monocyte percentage | 6.35 | 6.31 | 5388 | 160 | 0.95356 |
| Hemolysis index | 27.96 | 27.86 | 2463 | 126 | 0.98798 |

Polys, Polymorphonuclear Cells; WBC, White Blood Cell Count; RDW-SD, Blood Red Cell Distribution Width - Standard Deviation; MCHC, Blood Mean Corpuscular Hemoglobin Concentration; LDH, Blood Lactate Dehydrogenase; pO2, Blood Partial Pressure of Oxygen; PTT, Partial Thromboplastin Time.

### Feature Optimization to Maximize Predictive Performance for ICU Survival

Building on earlier findings, the Strict Survival Cohort (Survival_S) was constructed to predict ICU survival using a multimodal feature set. This cohort included a total of 5795 ICU encounters from MIMIC-IV with available CSF data and CSF culture results (Figure S1). LASSO-selected structured features included age, blood urea nitrogen and blood albumin levels that showed differential levels between survivor and non-survivor groups (Figure 3A, Tables 2 & S2). A fine-tuned BioBERT model was adapted to the proposed unstructured clinical text data, with tokenization optimized for Chief Complaint (CC), ICD entries (text format), and other clinical text features (Figure S3). The final input set integrated these text features with structured variables and CSF culture results. Three multimodal neural network architectures were evaluated: multimodal CNN (mCNN), multimodal FCNN (mFCNN), and multimodal LSTM (mLSTM) (Figure 3B). Among them, the mCNN model achieved the highest predictive performance (AUROC = 0.932) and was selected for downstream evaluation (Figure 3C).

**Figure 3.**
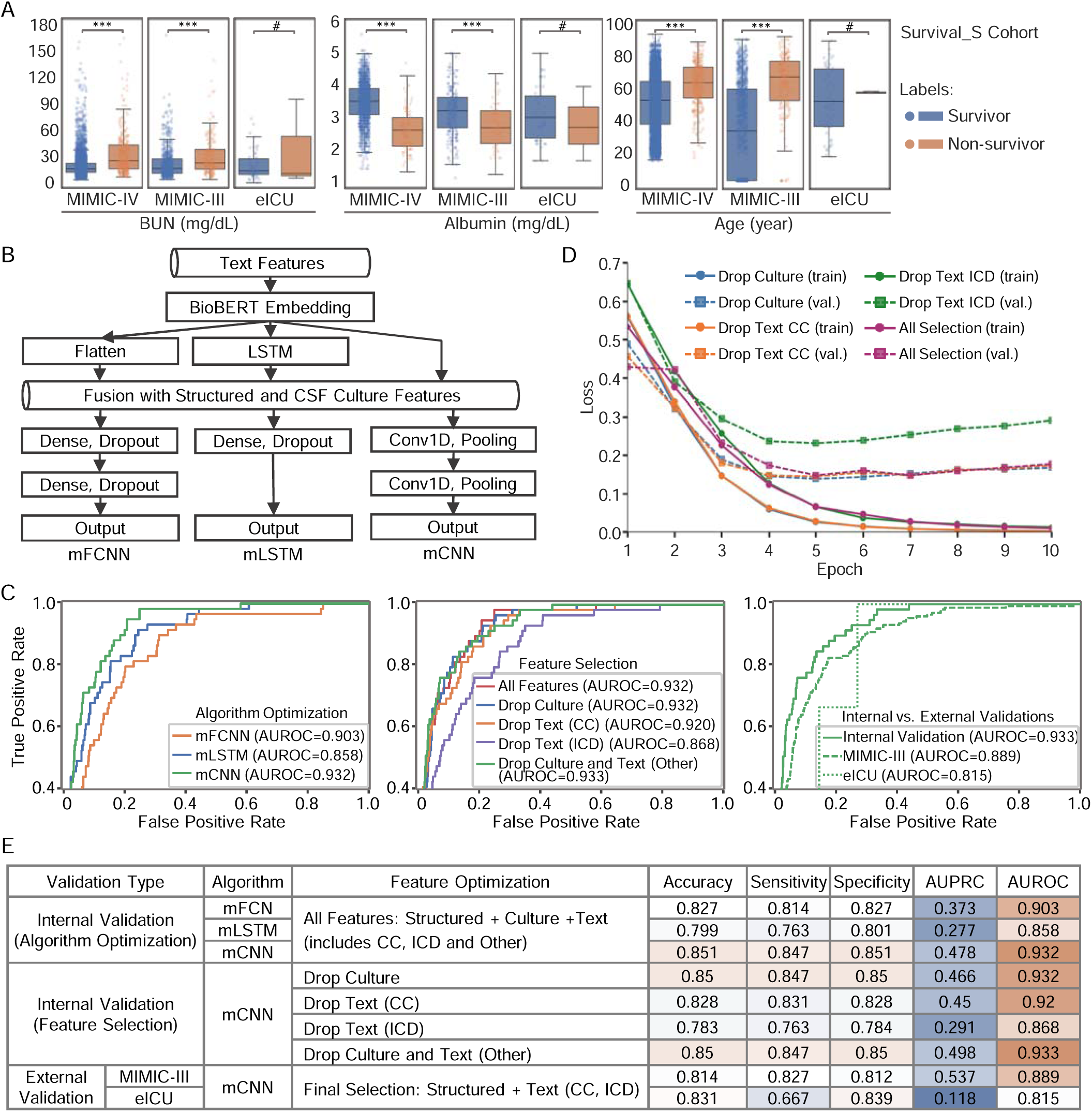
Model Development and Feature Optimization for Predicting ICU Survival. A. Three representative structured features in Survival_S cohort are plotted and compared between survivor and non-survivor groups across the MIMIC-IV, MIMIC-III, and eICU databases. B. Schematic diagram of three multimodal neural network architectures (mFCN, mLSTM, mCNN) that integrate structured features, CSF culture results, and text features processed by BioBERT. C. Receiver operating characteristic (ROC) curves showing internal model performance on the MIMIC-IV dataset for three algorithms (CNN, FCNN, RF) and under different feature selection settings. External validation performance of the optimized FCNN model is also shown for the MIMIC-III and eICU datasets. D. Training and validation loss curves over training epochs for mCNN models using full feature set with the exclusion of culture, chief complaint (CC), or ICD descriptions (in free-text format) features. E. Summary table of performance metrics across internal and external validation datasets using various combinations of structured features, CSF culture results, and text features. The text feature set includes CC, ICD (free-text format), and Other clinical text fields including illness history, medical history, allergies, and family history. The final selected feature set includes structured features along with two text features: ICD (free-text format) and chief complaint (CC). BUN: Blood Urea Nitrogen.

To evaluate the contribution of individual feature types, we conducted ablation tests during validation by removing selected feature inputs. Specifically, CSF culture results, CC, and ICD entries were individually or jointly excluded. Removing CSF culture alone, or in combination with other text data, did not impact validation loss or performance (AUROC remained 0.932) (Figure 3D & 3E). However, dropping CC or ICD alone reduced performance to AUROC = 0.920 and 0.868, respectively. Consistently, training and validation loss curves showed a noticeable increase in loss when ICD features were removed, while dropping CSF culture or other text features (e.g., CC) did not produce a similar increase. Independent validations in MIMIC-III (n = 1376) and eICU (n = 59) datasets yielded similar results, confirming the generalizability of these findings across diverse cohorts. These results underscore the predictive importance of unstructured text data, particularly ICD feature, in modeling ICU survival. In contrast, the CSF culture feature did not significantly impact model performance, likely because its predictive value is captured by structured lab test results already included in the model. As shown in the previous section, structured features alone were sufficient to accurately predict refined CSF culture results, suggesting redundancy and reduced incremental value from including the raw culture status.

**Table 2.**
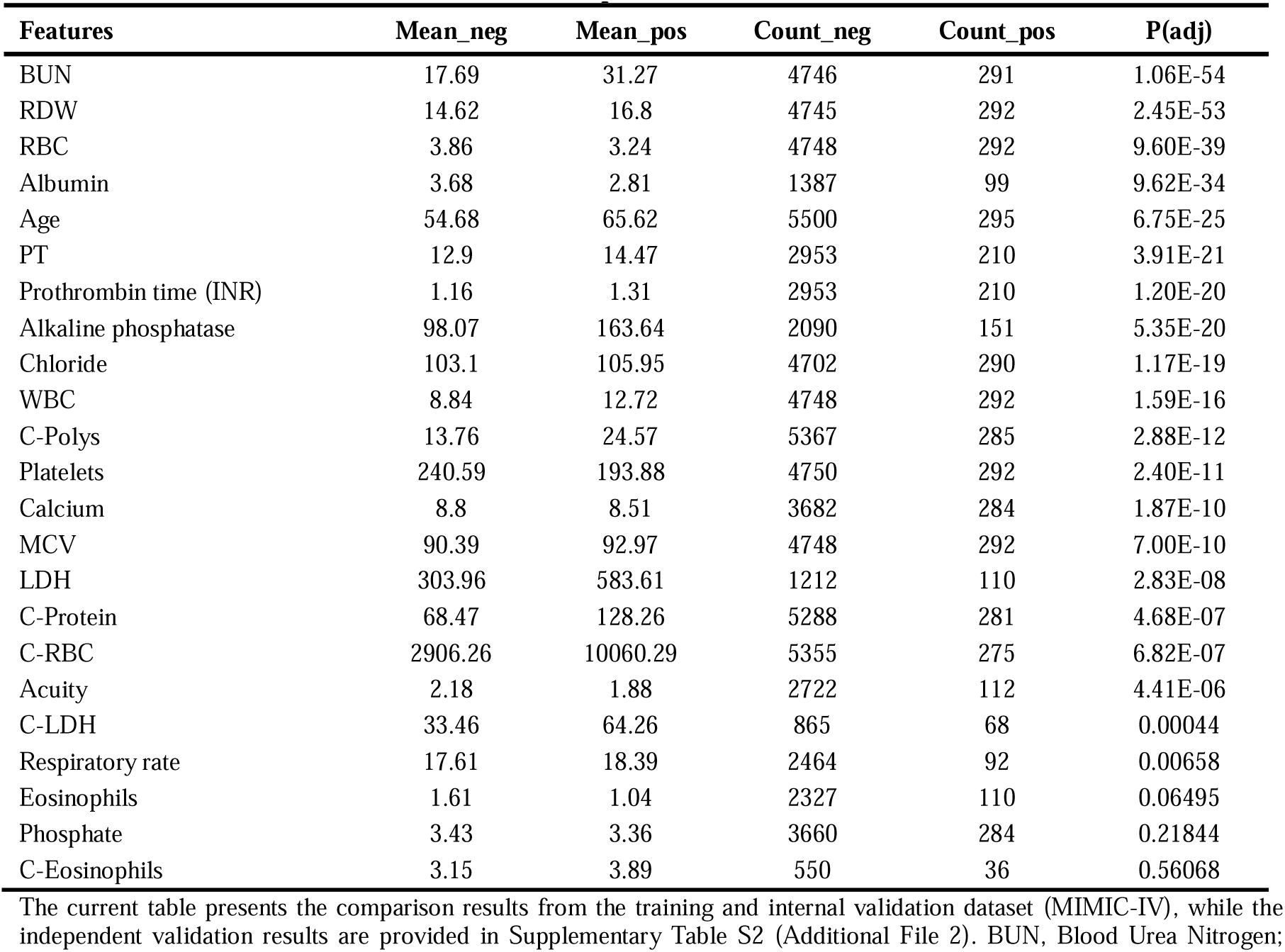
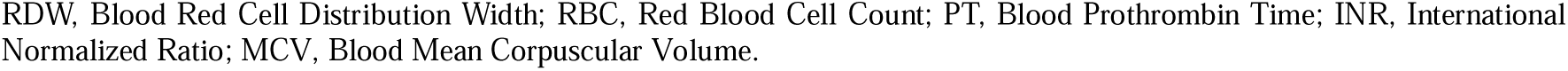
Baseline Characteristics and Mean Comparisons of the Survival_S Cohort (MIMIC-IV)

### Evaluation of NLP Contribution to ICU Survival Prediction

To further evaluate the predictive power of features identified in the Survival_S cohort and assess the specific contribution of NLP-based algorithms, we constructed a larger ICU survival cohort. By removing the requirement for CSF culture data, we expanded the eligible population to 431231 ICU admissions. This Lenient Survival Cohort (Survival_L) extended the previous work by relaxing inclusion criteria and eliminating culture features. It integrated structured variables adapted from a previously published ICU survival model by Lim et al.[8], along with free-text features identified through feature optimization in the Survival_S cohort. Most of the structured variables showed significant differences between survivors and non-survivors (Figure 4A, Tables 3 & S3). For unstructured data, top-ranked token features as converted by the BioBERT model included terms such as “fracture,” “pneumonia,” and “abnormal” from ICD fields, and “bleeding,” “weakness,” and “chest” from CC entries, which were significantly enriched in the survivor group (Figure 4B).

**Figure 4.**
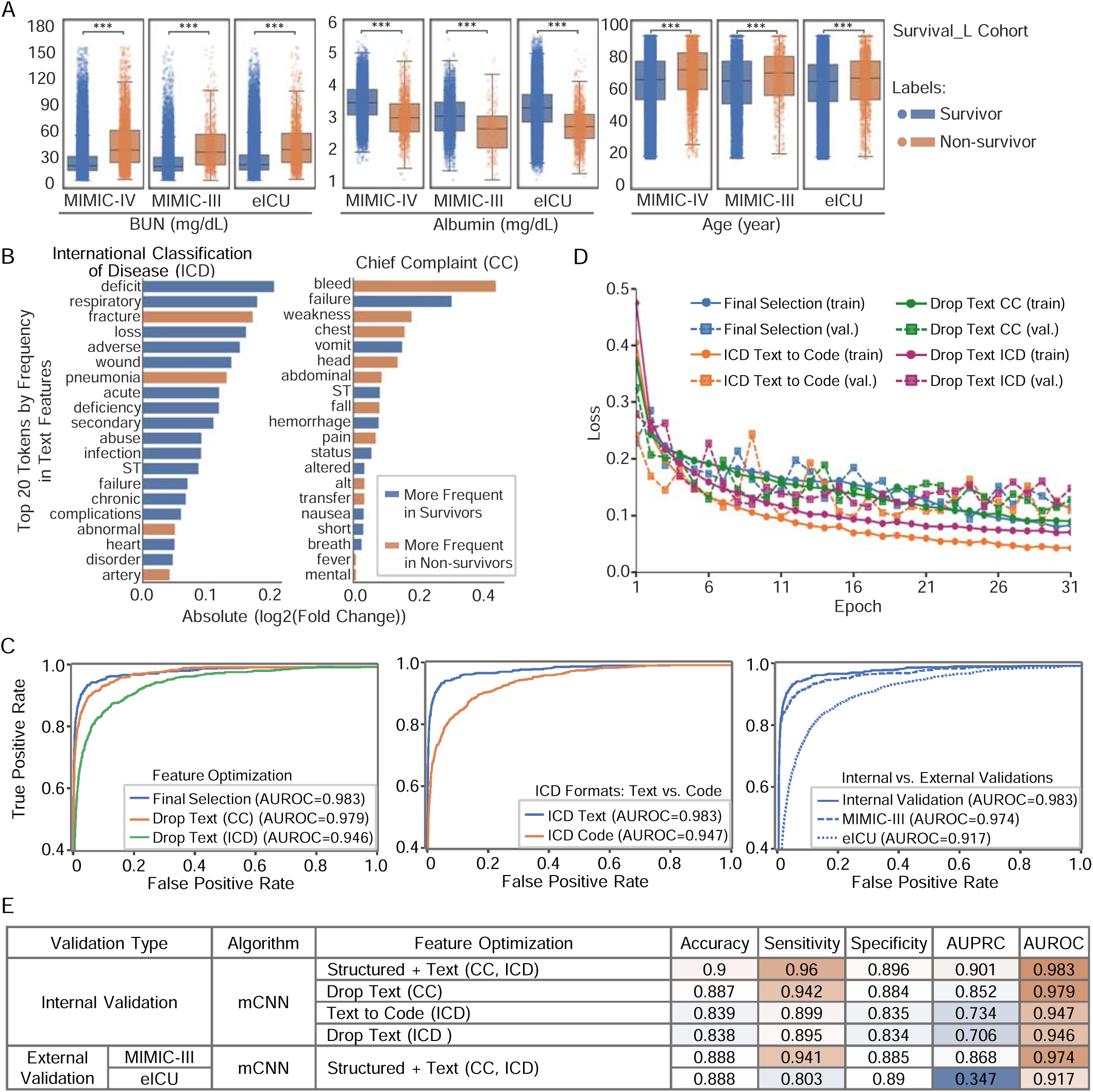
Model Evaluation for Predicting ICU Survival in an Extended Cohort and Assessing the Contribution of NLP-Based Text Features. A. Three representative structured features from the Survival_L cohort are plotted and compared between survivor and non-survivor groups across the MIMIC-IV, MIMIC-III, and eICU databases. B. Top 20 token words from the ICD and chief complaint (CC) fields that are enriched in survivor versus non-survivor groups, ranked by absolute log2 fold-change. C. Receiver operating characteristic (ROC) curves showing internal model performance on the MIMIC-IV dataset for feature sets comparing ICD text versus ICD code, and with exclusion of either CC or ICD. External validation performance of the optimized FCNN model is shown for the MIMIC-III and eICU datasets. D. Training and validation loss curves over training epochs for mCNN models using the final selected feature set, and after excluding specific features such as chief complaint (CC), ICD (free-text), or ICD with free-text converted to code format. E. Summary table of performance metrics across internal and external validation datasets using the final selected feature set with exclusion of chief complaint (CC), ICD (free-text), or ICD with free-text converted to code format.

The previously optimized mCNN model was applied to Survival_L to assess its generalizability and showed higher performance than Survival_S (AUROC = 0.983) (Figure 4C). Consistent with observations from the Survival_S cohort, excluding either CC or ICD text features individually resulted in a reduction in model performance (Figure 4D). Compared to the full model using all selected features, exclusion of CC reduced AUROC from 0.983 to 0.979, and exclusion of ICD reduced AUROC further to 0.946 (Figure 4E). Interestingly, when ICD was retained but converted from free text to code format, thus bypassing the need for BioBERT, the model’s AUROC still dropped to 0.947. These results confirm that the model maintained high performance in this large cohort (AUROC = 0.983, AUPRC = 0.901), and highlight the importance of using ICD in its original free-text form as processed by NLP algorithms to preserve predictive accuracy. Independent evaluation on external datasets further supported the robustness of the model. Validation in MIMIC-III (n = 26,836) and eICU (n = 200,895) resulted in AUROCs of 0.974 and 0.917, respectively, demonstrating consistent performance and strong generalizability across distinct ICU populations.

**Table 3.**
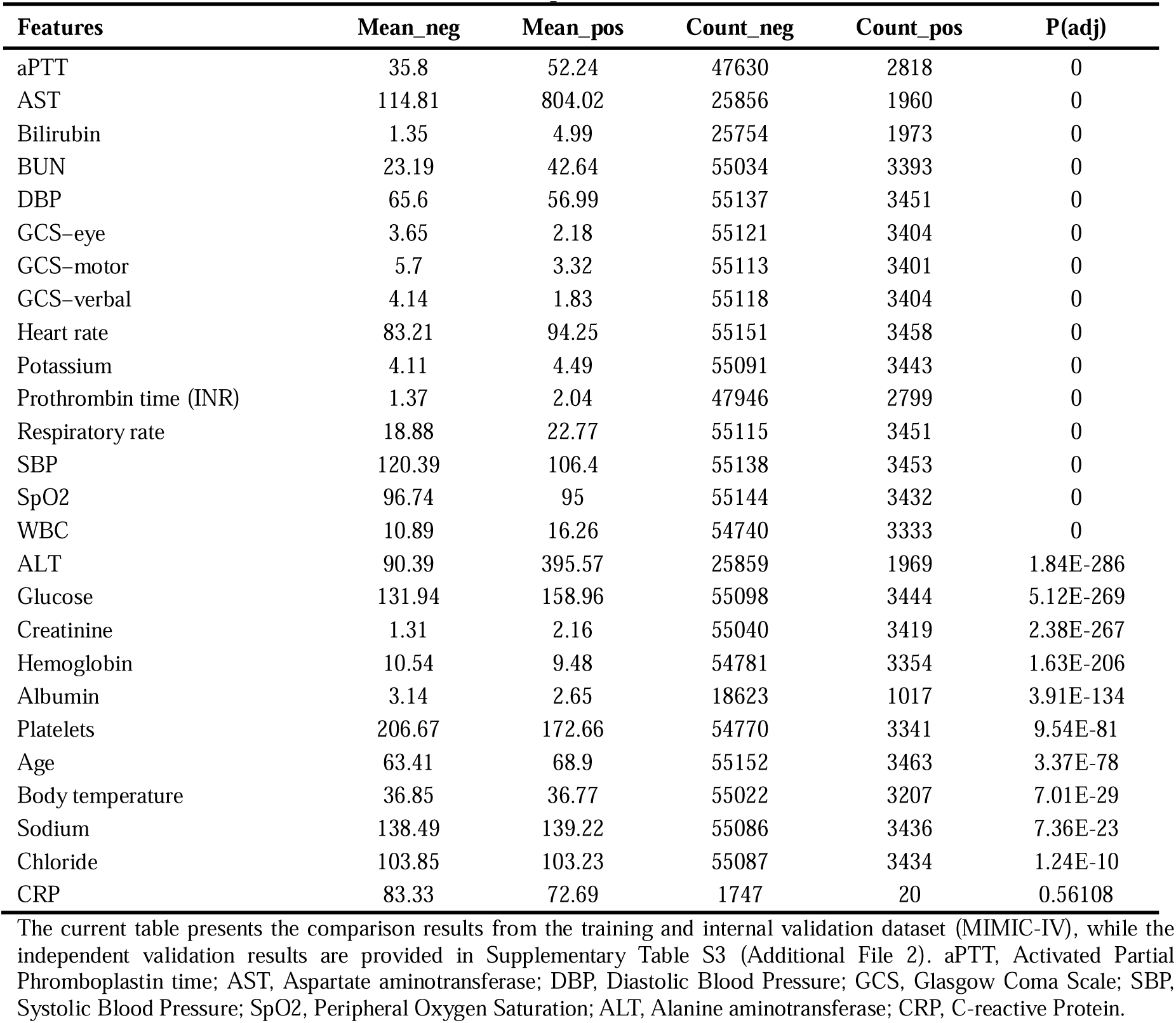
Baseline Characteristics and Mean Comparisons of the Survival_L Cohort (MIMIC-IV)

| Features | Mean_neg | Mean_pos | Count_neg | Count_pos | P(adj) |
| --- | --- | --- | --- | --- | --- |
| aPTT | 35.8 | 52.24 | 47630 | 2818 | 0 |
| AST | 114.81 | 804.02 | 25856 | 1960 | 0 |
| Bilirubin | 1.35 | 4.99 | 25754 | 1973 | 0 |
| BUN | 23.19 | 42.64 | 55034 | 3393 | 0 |
| DBP | 65.6 | 56.99 | 55137 | 3451 | 0 |
| GCS-eye | 3.65 | 2.18 | 55121 | 3404 | 0 |
| GCS-motor | 5.7 | 3.32 | 55113 | 3401 | 0 |
| GCS-verbal | 4.14 | 1.83 | 55118 | 3404 | 0 |
| Heart rate | 83.21 | 94.25 | 55151 | 3458 | 0 |
| Potassium | 4.11 | 4.49 | 55091 | 3443 | 0 |
| Prothrombin time (INR) | 1.37 | 2.04 | 47946 | 2799 | 0 |
| Respiratory rate | 18.88 | 22.77 | 55115 | 3451 | 0 |
| SBP | 120.39 | 106.4 | 55138 | 3453 | 0 |
| SpO2 | 96.74 | 95 | 55144 | 3432 | 0 |
| WBC | 10.89 | 16.26 | 54740 | 3333 | 0 |
| ALT | 90.39 | 395.57 | 25859 | 1969 | 1.84E-286 |
| Glucose | 131.94 | 158.96 | 55098 | 3444 | 5.12E-269 |
| Creatinine | 1.31 | 2.16 | 55040 | 3419 | 2.38E-267 |
| Hemoglobin | 10.54 | 9.48 | 54781 | 3354 | 1.63E-206 |
| Albumin | 3.14 | 2.65 | 18623 | 1017 | 3.91E-134 |
| Platelets | 206.67 | 172.66 | 54770 | 3341 | 9.54E-81 |
| Age | 63.41 | 68.9 | 55152 | 3463 | 3.37E-78 |
| Body temperature | 36.85 | 36.77 | 55022 | 3207 | 7.01E-29 |
| Sodium | 138.49 | 139.22 | 55086 | 3436 | 7.36E-23 |
| Chloride | 103.85 | 103.23 | 55087 | 3434 | 1.24E-10 |
| CRP | 83.33 | 72.69 | 1747 | 20 | 0.56108 |

## Discussion

This study presents a stepwise machine learning framework aimed at improving outcome prediction for critically ill patients by leveraging both structured clinical data and unstructured text information. While CSF culture remains the diagnostic gold standard for bacterial meningitis, we first explored whether structured laboratory features could support culture result prediction. Although not intended to replace microbiological diagnostics, this preliminary model demonstrated that routinely collected data may help flag high-probability cases, potentially assisting early decision-making in settings where culture turnaround is delayed or results are inconclusive.

Building on this foundation, we then focused on predicting ICU survival outcomes using a multimodal deep learning approach. The Survival_S cohort integrated structured features, CSF culture data, and unstructured clinical text (such as chief complaints and ICD entries), enabling a more comprehensive risk profile. Finally, the Survival_L cohort extended this approach to a larger, more inclusive patient population by removing the CSF culture requirement and testing the model’s generalizability. Across these cohorts, the mCNN model consistently demonstrated strong predictive performance, and ablation experiments confirmed the value of NLP-transformed text features in improving survival prediction.

A standard CSF culture typically takes 1 to 3 days to grow and identify bacteria under lab conditions[26]. Clinicians are increasingly relying on molecular tests like Polymerase Chain Reaction (PCR) for identifying pathogens due to PCR’s speed and accuracy in detecting bacterial DNA[27, 28]. Observing the limitations of culture results alone, we modified the machine learning input data in our BM diagnosis model training to use refined culture labels that integrate additional clinical evidence. Here our study demonstrated the effectiveness of selective algorithms in utilizing regular blood and CSF test result and accurately predicting positivity of CSF culture results, achieving an AUROC of 0.853 and 0.849 in the validation datasets. This result shows promise of the CNN model as a valuable tool in assisting clinical decision-making with a critical advantage by providing results instantly, bypassing the typical 24 to 72-hour delay as required for traditional CSF culture.

MIMIC-IV, as an evolution of MIMIC-III, has been widely used for developing and testing complex modeling machine learning models in healthcare research[15, 29]. The key improvements of MIMIC-IV include enhanced details on diagnoses and procedures using ICD-10 codes (where MIMIC-III used ICD-9 codes), and more refined dataset with better support for unstructured data processing through tools like NLP. MIMIC-IV has been extensively employed in mortality prediction and survival analysis. Classical machine learning models such as XGBoost and logistic regression often achieve robust AUROC scores around 0.80 to 0.85[30–32]. This level of performance makes these models strong candidates for applications in clinical prediction tasks, especially when interpretability and computational efficiency are key requirements. Taking advantage of the updated and additional data integrated in MIMIC-IV, the hypothesis of this study was that multimodal machine learning model can deliver precise survival prediction and personalized patient management. BERT and its model variants (BioBERT, Bio_ClinicalBERT etc.) have been widely used to extract textual features and their relationships from free-text biomedical data[10, 11, 33]. BioBERT, an extension of BERT, was pre-trained on PubMed abstracts and full-text articles from PubMed Central, both of which are databases rich in biomedical literature[10, 11, 33]. Bio_ClinicalBERT, on the other hand, was trained on clinical notes from the MIMIC-III dataset, making it more specialized for clinical text. However, its application in large urgent care datasets remains limited. In this study, independent validation was performed using data from the MIMIC-III database. To minimize the risk of information leakage between training and validation datasets, we selected BioBERT rather than Bio_ClinicalBERT for text embedding. Interestingly, when comparing outputs from both models on clinical text such as chief complaints and ICD entries, we observed that the resulting feature vectors were identical. This suggests that, despite differences in their pretraining corpora, both models can generate consistent representations for certain types of structured clinical text. A prior study has demonstrated the utility of BioBERT in mortality prediction by encoding time-series and clinical text data from MIMIC-III[34]. Consistent with these findings, our results showed that integrating BioBERT to process ICD text entries in the MIMIC-IV dataset substantially improved the performance of deep learning multi-models. Notably, encoding ICD entries in their original free-text form using NLP significantly outperformed models that relied on directly inputting structured ICD codes. The integration led to our best-performing model achieving an AUROC greater than 0.970 in both internal and external validations (MIMIC-IV and MIMIC-III). These findings underscore the value of incorporating advanced NLP techniques to extract meaningful information from clinical narratives and enhance predictive modeling in critical care

Together, this study demonstrates the potential of an integrated machine learning–based framework to support patient management and enhance clinical decision-making by leveraging natural language processing (NLP) for clinical text data such as ICD entries. Applying NLP to transform free-text ICD data significantly increases its utility in predictive model construction, unlocking greater accuracy and timeliness in the clinical management of CNS infections and critically ill ICU patients. By combining structured and unstructured data processed through advanced machine learning techniques, this multimodal approach offers a precise and scalable solution to improve risk stratification and care prioritization in critical care settings. For instance, such a system could include tools like a Clinical Decision Support System to would enable clinicians to proactively identify patients at high risk of adverse outcomes and guide early interventions[35].

A potential limitation of both the diagnostic and survival prediction models is their relatively lower AUPCR compared to AUROC, indicating a higher rate of false positives and reduced precision in identifying true positive cases. In the diagnostic setting for BM, false positives may lead to unnecessary or prolonged antibiotic use. However, this is considered a minor and acceptable risk, as clinical guidelines recommend initiating antibiotic treatment within one hour of suspected BM presentation[36, 37], and over 90% of suspected cases receive empiric antibiotics before or immediately after lumbar puncture[38]. in the ICU context, a conservative prediction bias may be acceptable, especially for high-risk conditions, where early intervention is critical. False positives in this setting may lead to closer monitoring or precautionary treatment, which are generally low-risk actions but could contribute to resource strain in overwhelmed ICUs. Notably, the AUPRC in the expanded Survival_L cohort showed substantial improvement, achieving 0.901 in internal validation and 0.868 on external validation with the MIMIC-III dataset. These values indicate strong precision in predicting ICU mortality in large, diverse patient populations.

## Conclusion

This study demonstrates the utility of multimodal machine learning models for predicting CSF culture results and ICU survival outcomes. Integrating structured data with NLP-processed free-text inputs, especially ICD entries and chief complaints, significantly improved predictive performance. These findings highlight the value of unstructured clinical text in enhancing risk stratification and support the development of real-time decision tools for ICU care.

## Data Availability

The data utilized in this study are derived from the MIMIC-III (https://doi.org/10.13026/8a4q-w170), MIMIC-IV (https://doi.org/10.13026/a3wn-hq05) and eICU (https://doi.org/10.13026/4mxk-na84) critical care databases, which are accessible to researchers who fulfill the credentialing requirements set by the data custodians. Due to these restrictions, the raw data are not publicly available. However, upon obtaining the necessary permissions from the MIMIC data providers, the data can be accessed for replication or further study.
The complete set of code for implementing the machine learning models, including those predicting CSF culture results and ICU outcomes, is openly available in our GitHub repository: https://github.com/AaronChen007/CNS_Model_Code.

## Abbreviations

AUPRC: Area Under the Precision-Recall Curve
AUROC: Area Under the Receiver Operating Characteristic Curve
BERT: Bidirectional Encoder Representations from Transformers
BioBERT: BERT pretrained on biomedical data
BM: Bacterial meningitis
CC: Chief Complaint
CDSS: Clinical Decision Support System
CNN: Convolutional Neural Network
CSF: Cerebrospinal fluid
FCN: Fully connected neural network
ICD: International Classification of Diseases
ICU: Intensive Care Unit
LASSO: Least Absolute Shrinkage and Selection Operator
LSTM: Long short-term memory
MIMIC: Medical Information Mart for Intensive Care
NLP: Natural Language Processing
PCR: Polymerase Chain Reaction
RF: Random Forest

## Supplementary Information

**Additional file 1: Figure S1.** Overview of Data Extraction, Model Construction, and Validation Workflow. **Figure S2.** Feature Selection with LASSO. **Figure S3.** Cumulative probability distributions of token counts in unstructured clinical text processed by BioBERT.

**Additional file 2: Table S1.** Baseline Characteristics and Mean Comparisons of the Culture Cohort. **Table S2.** Baseline Characteristics and Mean Comparisons of the Survival_S Cohort. **Table S3.** Baseline Characteristics and Mean Comparisons of the Survival_L Cohort. **Table S4.** Mapping of Feature Short Names to Full Descriptions.

## Acknowledgments

We extend our gratitude to the MIT Laboratory for Computational Physiology and the respective data curators for creating and maintaining the MIMIC-III, MIMIC-IV, and eICU critical care databases. We also acknowledge the patients whose data contributed to these datasets, enabling this research. Without their generous participation and the efforts of those involved in the MIMIC and eICU projects, our study would not have been possible.

## Author contributions

XZ designed the workflow, drafted initial analysis code, finalized the manuscript, and supervised all aspects of the study. RC and CY built the data-analysis pipeline, performed experiments, analyzed and illustrated the data, and prepared the initial manuscript draft. SZ contributed to data interpretation and assisted in data visualization. ZH, MS, WL, DY, SH and LB aided in experimental design and reviewed the manuscript.

## Funding

This work was supported by National Natural Science Foundation of China (81672627; 82071863).

## Availability of data and materials

The data utilized in this study are derived from the MIMIC-III (https://doi.org/10.13026/8a4q-w170), MIMIC-IV (https://doi.org/10.13026/a3wn-hq05) and eICU (https://doi.org/10.13026/4mxk-na84) critical care databases, which are accessible to researchers who fulfill the credentialing requirements set by the data custodians. Due to these restrictions, the raw data are not publicly available. However, upon obtaining the necessary permissions from the MIMIC and eICU data providers, the data can be accessed for replication or further study.

The complete set of code for implementing the machine learning models, including those predicting CSF culture results and ICU outcomes, is openly available in our GitHub repository: https://github.com/AaronChen007/CNS_Model_Code.

## Declarations

### Ethics approval and consent to participate

The research utilized the MIMIC and eICU database, which was initially approved for use by the Institutional Review Boards of Beth Israel Deaconess Medical Center and the Massachusetts Institute of Technology. Since MIMIC and eICU are publicly available, de-identified datasets, individual patient consent was not required. All analyses and research activities strictly adhered to the MIMIC and eICU data access agreement and data usage guidelines.

## Consent for publication

All authors have read the manuscript and are consentaneous for publication.

## Competing interests

The authors have declared that no conflict of interest exists.

## Notes

### Competing Interest Statement

The authors have declared no competing interest.

### Author Declarations

The research utilized the MIMIC database (physionet.org/content/mimiciv/3.1/), which was initially approved for use by the Institutional Review Boards of Beth Israel Deaconess Medical Center and the Massachusetts Institute of Technology. Since MIMIC is a publicly available, de-identified dataset, individual patient consent was not required. All analyses and research activities strictly adhered to the MIMIC data access agreement and data usage guidelines.

### Summary of Updates

In this revised version of the manuscript, we have incorporated a few updates to strengthen the study. Most notably, we added a new ICU survival analysis cohort, referred to as the Lenient ICU Survival Cohort (Survival_L), which expands upon the original Strict ICU Survival Cohort by removing the cerebrospinal fluid (CSF) culture requirement, thereby including a broader and more diverse patient population. This addition allows for a more comprehensive evaluation of model performance and generalizability across varied ICU populations.

