## Additional File 1 for "Contextualized Biomedical Language Processing Enhances ICU Survival Prediction"

**Appendix**

**Figure S1.** Overview of Data Extraction, Model Construction, and Validation Workflow.

**Figure S2.** Feature Selection with LASSO.

**Figure S3.** Cumulative probability distributions of token counts in unstructured clinical text processed by BioBERT.

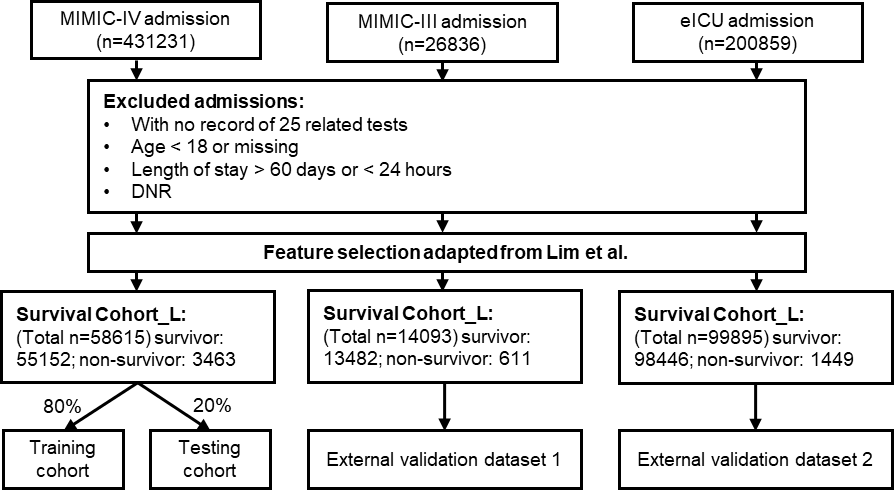

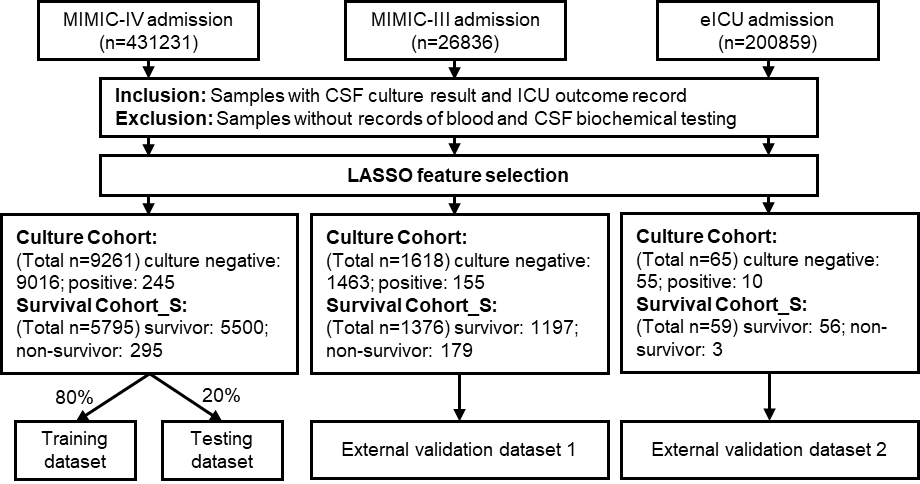

**B**

**A**

**Figure S1.** Overview of Data Extraction, Model Construction, and Validation Workflow.
(A) Development of Culture and ICU Mortality Prediction Models. Data were retrospectively extracted from the MIMIC‐IV, MIMIC‐III subset, and eICU databases. Cases with CSF culture results and ICU outcome records were included, whereas samples lacking both blood and CSF biochemical testing were excluded. This procedure generated separate Culture and Survival Datasets for each database. The MIMIC‐IV datasets were partitioned into training (80%) and testing (20%) cohorts, with the MIMIC‐III subset and eICU datasets serving as external validation cohorts. Separate models were trained on the MIMIC‐IV training cohorts for predicting CSF culture status and ICU mortality, and subsequently validated on both internal and external cohorts.

(B) From admission records of MIMIC‐IV, MIMIC‐III subset, and eICU databases, cases were excluded if patients lacked data for 25 relevant clinical tests, were younger than 18 years or had missing age information, had ICU stays longer than 60 days or shorter than 24 hours, or were designated as 'Do Not Resuscitate' (DNR). The resulting survival datasets comprised 26 structured variables (nine vital signs, 16 laboratory tests and age) and two unstructured variables (chief complaint and ICD text). The MIMIC‐IV dataset was divided into training (80%) and testing (20%) cohorts, while the MIMIC‐III subset and eICU cohorts served as external validation datasets.

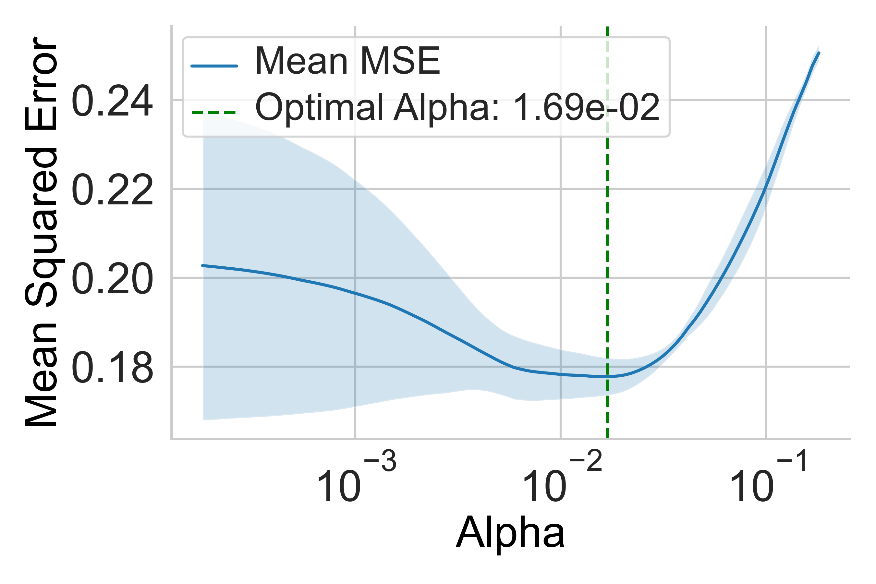

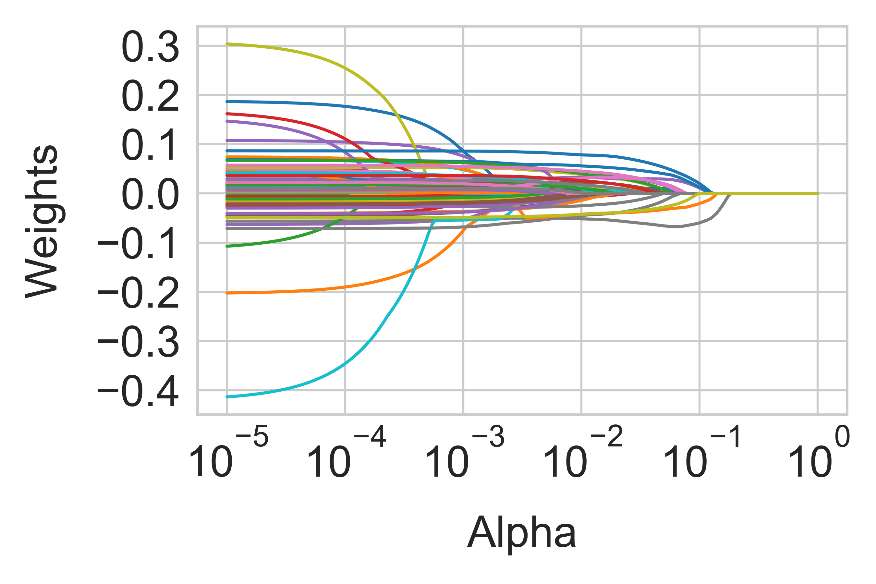

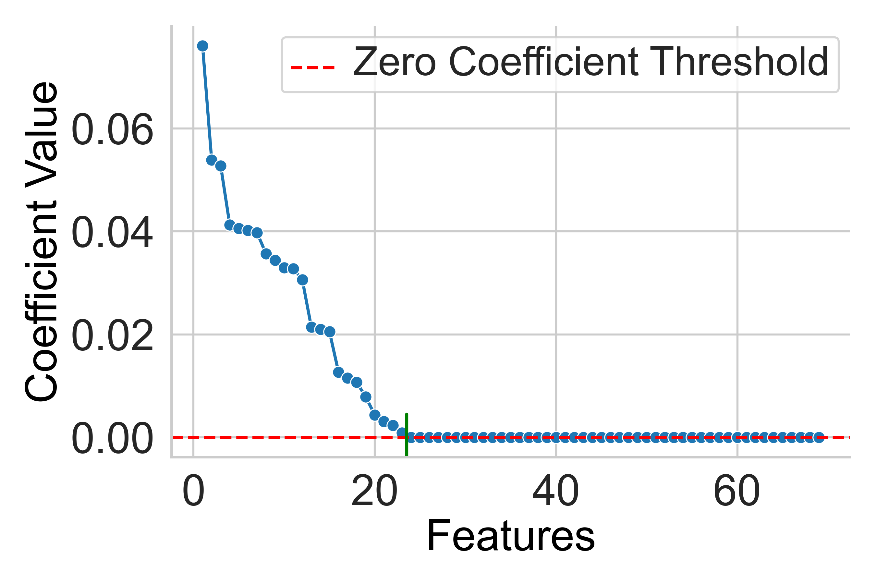

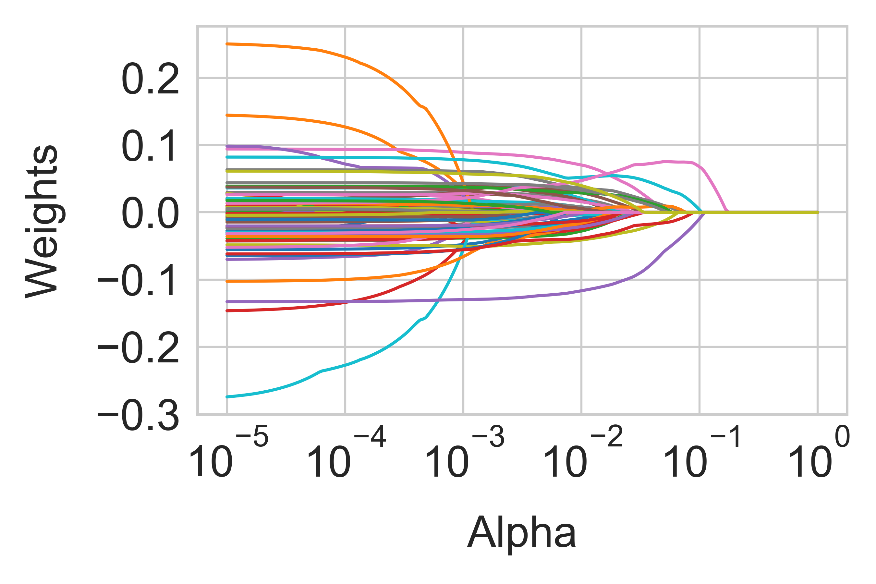

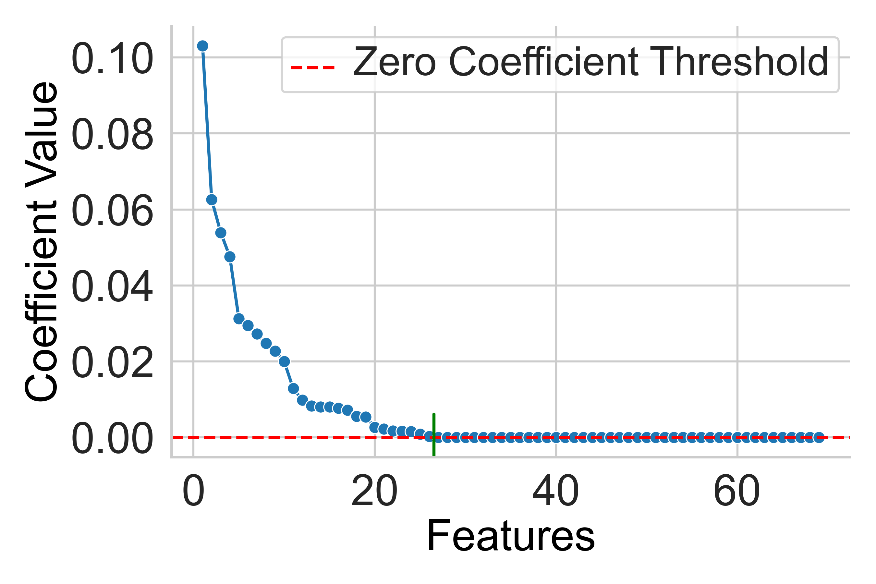

**A**

**B**

**C**

**D**

**E**

**F**

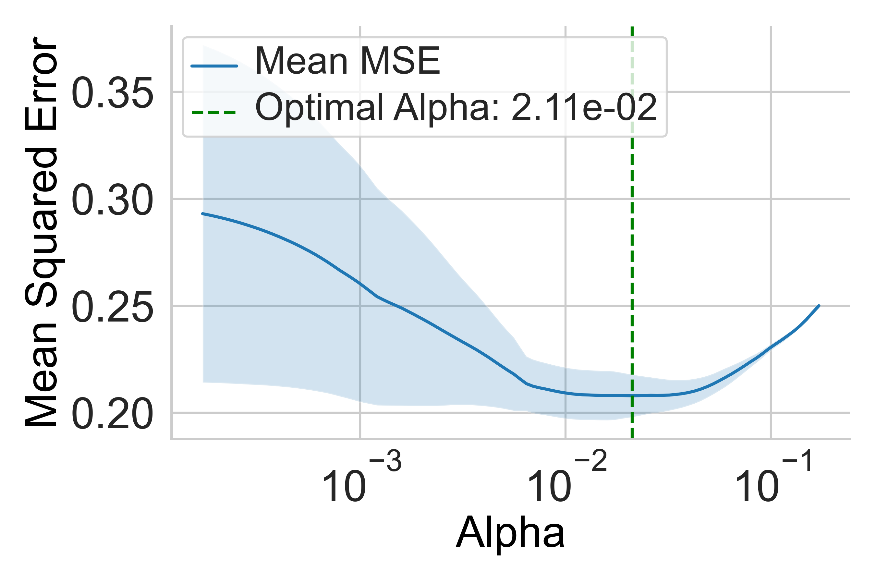

**Figure S2.** Feature Selection with LASSO. (A-C) show the feature selection for the Culture Dataset, and (D-F) depict the process for the Survival Dataset. (A & D) Display the coefficients of all features, ordered by their absolute values. The red dashed line indicates the zero-coefficient threshold. (B & E) Illustrate the relationship between the regularization parameter alpha and model coefficients, with the x-axis on a logarithmic scale. Green dashed lines in (C & F) mark the optimal alpha value, determined through cross-validation to minimize the MSE.

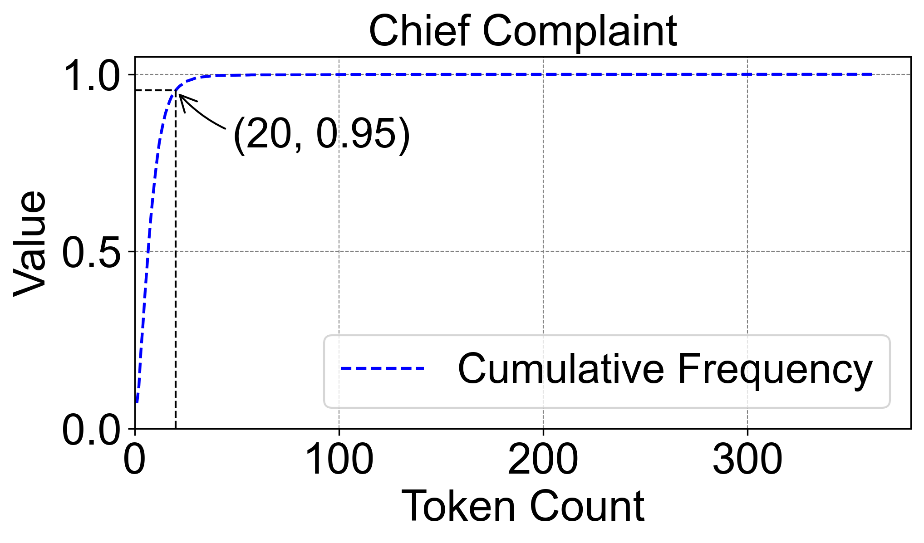

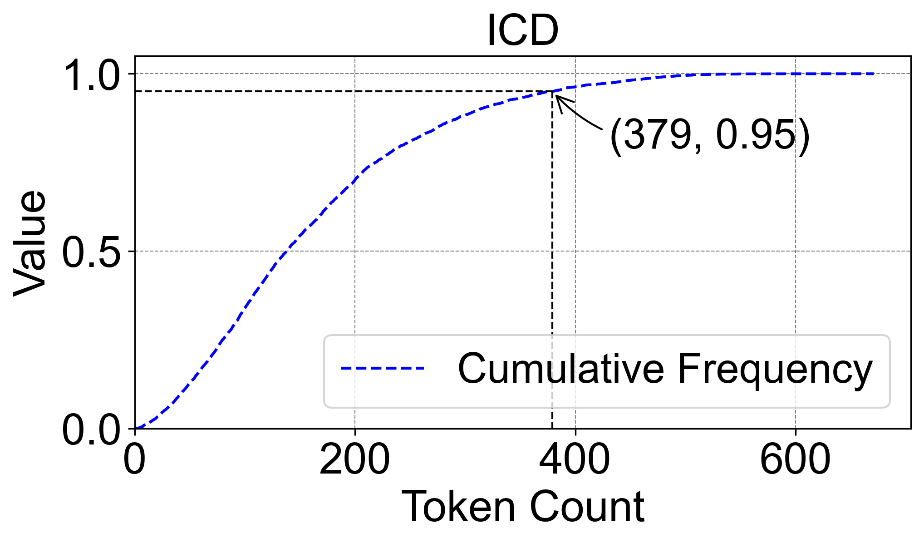

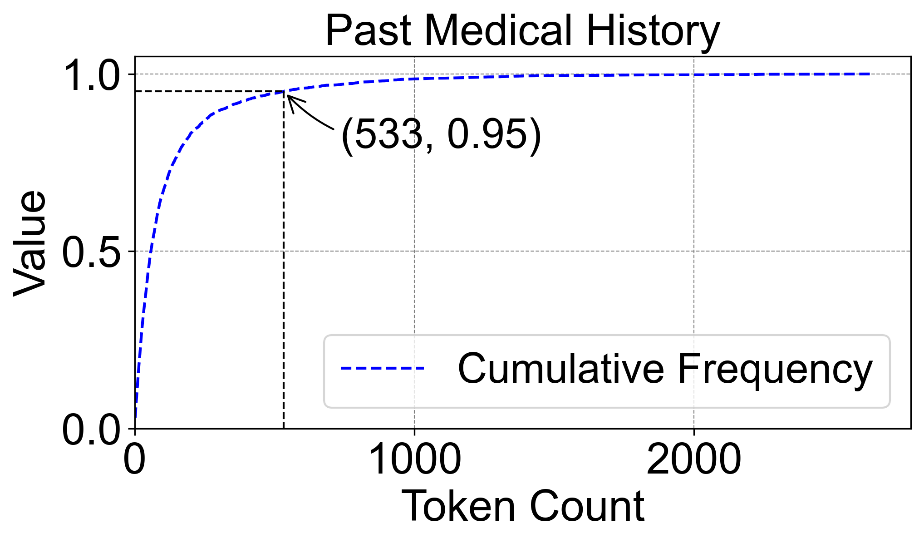

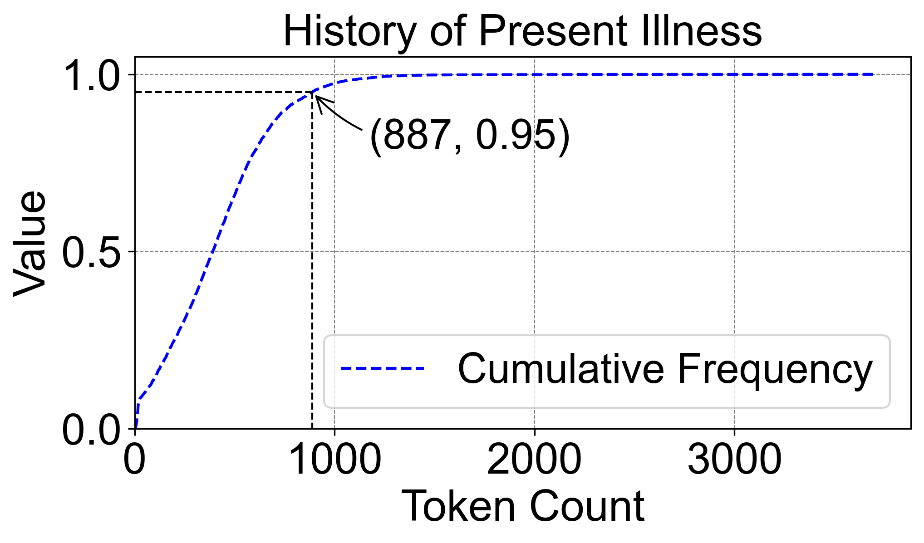

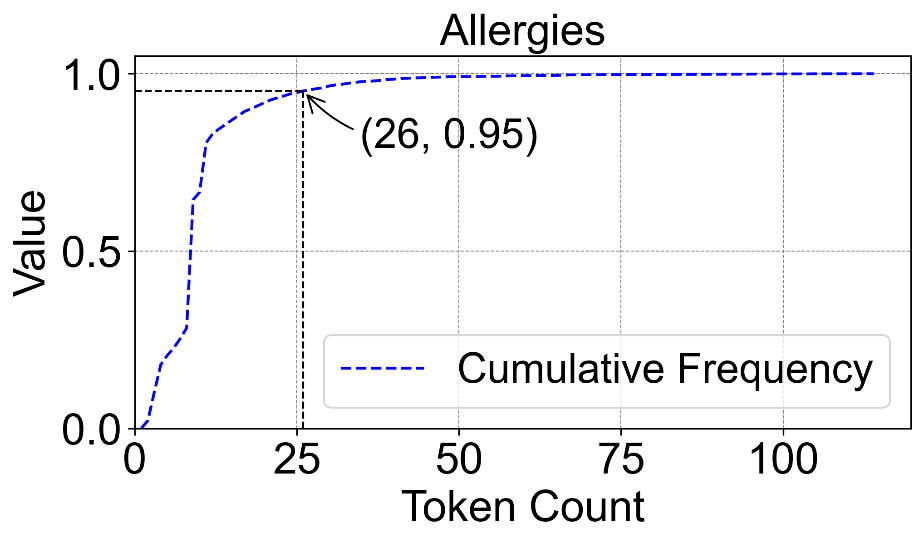

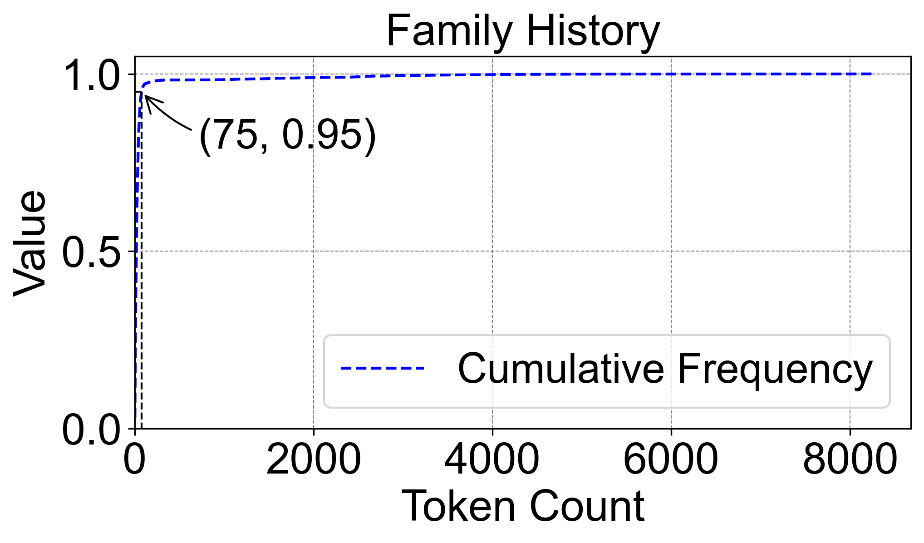

**A**

**B**

**C**

**E**

**F**

**D**

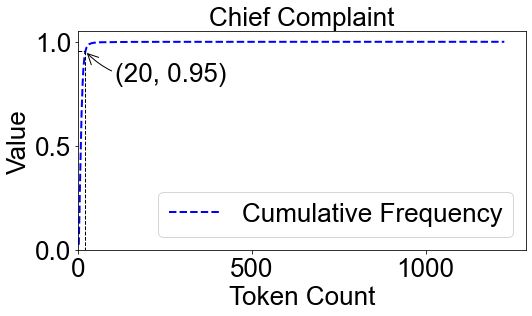

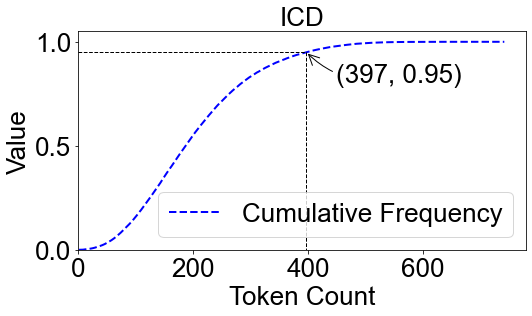

**G**

**H**

**Figure S3.** Cumulative probability distributions of token counts in unstructured clinical text processed by BioBERT. Panels A–F display the cumulative probability plots of token counts derived from unstructured text data in the Survival_S cohort, while panels G and H correspond to the Survival_L cohort. All unstructured data were tokenized using BioBERT to quantify the textual complexity and distribution of input lengths across cohorts.
